# Endoscopic liquid biopsies of gastric fluid in a large patient cohort reveal DNA content as a candidate tumor biomarker in gastric cancer

**DOI:** 10.1101/2023.02.14.23285919

**Authors:** Francine Carla Cadoná, Thais F. Bartelli, Adriane Graicer Pelosof, Claudia Zitron Sztokfisz, Adriana Passos Bueno, Luana Batista do Carmo dos Santos, Gabriela Pereira Branco, Gabriel Oliveira dos Santos, Warley Abreu Nunes, Fernanda Araújo Pintor, Laís Lie Senda de Abrantes, Alexandre Defelicibus, Luiz Gonzaga Vaz Coelho, Marcis Leja, Haejin In, Sharon Li, Howard Hochster, Felipe José Fernandez Coimbra, Rodrigo D. Drummond, Israel Tojal da Silva, Ravi J. Chokshi, Renata Pasqualini, Wadih Arap, Diana Noronha Nunes, Emmanuel Dias-Neto

**Author notes:** Department of Clinical Cancer Prevention, The University of Texas M. D. Anderson Cancer Center, Houston, Texas, USA. Address correspondence to: Wadih Arap or Emmanuel Dias-Neto, Rutgers Cancer Institute, 250 South Orange Avenue, Newark, New Jersey 07101, USA. (WA) or (ED-N).

## Abstract

**Background:** Human gastric cancer remains a diagnostic and therapeutic challenge worldwide. Improved prognostic biomarkers may support treatment planning, including surgery, neoadjuvant and adjuvant therapies. We offer a new liquid biopsy approach, integrated with esophagogastroduodenoscopy (EGD) to evaluate gastric fluid recovered from a large patient cohort (n=1,056 patients), demonstrating the value of a simple DNA concentration analysis to provide diagnostic support and prognosis in gastric cancer.

**Methods:** We performed an exploratory study to evaluate gastric fluid DNA (gfDNA) concentration in patients with normal gastric mucosa or diagnosed with peptic diseases, preneoplastic conditions, or cancer. Key variables including sex, gastric fluid pH, use of proton-pump inhibitors, tumor subtype, clinical stage, and disease outcomes were considered.

**Results:** gfDNA concentrations were significantly increased in gastric cancer versus all groups (mean 26.86 ng/µL; 95% CI: 20.05 to 33.79; p=3.61e^-12^) and as compared to non-malignant controls (normal mucosa and peptic diseases; mean 10.77 ng/µL; 95% CI: 9.23 to 12.33; p=9.55e^-13^), and preneoplastic conditions (mean 10.10 ng/µL; 95% CI: 7.59 to 12.60; p=1.10e^-5^). gfDNA concentrations were higher in advanced tumors (T3; mean 25.66 ng/µL; 95% CI: 19.46 to 31.85) compared to early-stage disease (T2 and below; mean 15.12 ng/µL; 95% CI: 9.73 to 20.50; p=5.97e^-4^). Our findings also suggest gfDNA prognostic value for gastric cancer diagnosed subjects. In this subset, patients with gfDNA concentrations >1.28 ng/µL showed longer progression-free survival (p=0.009), which correlated with increased tumor-infiltrating immune cells (p=0.001), remaining significant after adjusting for tumor stage (p=0.014).

**Conclusions:** While high gfDNA indicates gastric cancer in a general population and may contribute to disease management, its diagnostic clues should be integrated with further work up such as tissue biopsies, to ensure comprehensive and accurate clinical assessment. However, in patients with established gastric cancer, gfDNA is a potential prognostic marker, and subjects with high gfDNA content might paradoxically have better outcomes. The elevated presence of immune cell infiltrates—compared to those observed in peptic diseases and pre-neoplastic lesions (including their associated gastric cavity DNA shedding)—may serve as a critical missing link to resolve this empirical paradox.

## Introduction

Esophagogastroduodenoscopy (EGD) is routinely performed to work up vague digestive complaints and is used to biopsy suspicious lesions and ultimately diagnose gastric cancer, a tumor that ranks as the fifth in terms of incidence and mortality in the world, with current estimates of ∼660,000 annual deaths globally (Globocan, 2022). In areas of high-incidence EGD has been shown to contribute to a reduction in gastric cancer mortality (Hamashima et al., 2013; Matsumoto and Yoshida, 2014), and is also employed for presurgical gastric cancer staging and even as a treatment method (endoscopic resection) in early-stage disease (Kim and Jung, 2021), rendering it a universally used tool in this setting. During EGD, gastric fluids are collected and discarded, allowing a better evaluation of the gastric mucosa.

The study of DNA found in body-fluid samples (e.g., blood, urine, saliva, cerebrospinal fluid, peritoneal washes) can support the diagnosis and/or follow-up of cancer (Forshew et al., 2012). In a liquid biopsy approach, tumor-derived DNA permits the monitoring of specific mutations diluted in the fluids under investigation. As the gastric fluid is in direct contact with the stomach mucosa and gastric cancer lesions, liquid biopsies performed with gastric fluid are neither restricted to just the few selected areas sampled by tissue biopsy, nor by the minute amounts of tumor DNA that might be diluted in peripheral blood. Also, it may indeed allow a full representation of cells that reach the stomach, derived from the upper digestive tract, including immune cells, tumor cells and the microbiota. In a prior feasibility study, we showed that gastric fluid incidentally collected from gastric cancer patients during EGD contains tumor-derived DNA, allowing the capture of tumor mutations (Pizzi et al., 2019). We have now collected gastric fluids from a large patient cohort (n=1,056), obtained during diagnostic endoscopy, to investigate whether gastric fluid DNA (gfDNA) concentrations would vary according to disease diagnosis, including the presence of non-malignant, premalignant, or malignant lesions, gastric cancer subtype and stage, tumor location, and patient age or sex. Notably, we also evaluated whether gfDNA concentrations would have predictive and prognostic value on gastric cancer recurrence and patient outcomes.

## Methods

### Patients

All subjects enrolled in this study underwent diagnostic EGD at a single institution, the A. C. Camargo Cancer Center, São Paulo, Brazil. Inclusion criteria required subjects to be above 18 years old, with clinical indication for performing a diagnostic EGD examination. Samples and analyses were excluded if requested by the participants (even after the signature of a written informed consent), or if other unrelated diagnoses were found (e.g., previous gastrectomy or presence of other non-gastric cancer. Samples were collected at diagnosis, during standard EGD, from February 2016 to March 2021. EGDs were only performed after referral from an independent physician (i.e., not related to the study) to assess abdominal symptoms, under standard sedation. Samples were obtained after either patients or their guardians signed a written informed consent form approved by the Clinical Research Committee, the Ethics Research Committee, and the Institutional Review Board (IRB) (protocol #2134/15). Diagnostic and epidemiological data were collected as described (Bartelli et al., 2019).

### Gastric fluid collection, DNA extraction and quantification

As usual and customary for upper endoscopic examination, all subjects were fasting for 8-12 hours. At the beginning of the procedures, gastric fluids that are routinely removed to allow better examination of the mucosa, were collected in sterile plastic containers attached to the endoscope suction channel and kept on ice until pH measurement and neutralization. Samples were divided in aliquots and kept frozen at -20°C (each aliquot was thawed just once), until DNA was extracted from an equal volume of gastric fluid (800 µl) by proteinase-K digestion, followed by phenol-chloroform extraction and ethanol DNA precipitation. DNA was subsequently resuspended in 100 µl of sterile nuclease-free double-deionized water and quantified by Qubit fluorometry (Thermo Fisher).

### Quantification of human- versus bacteria-derived DNA in gastric fluids

For a representative fraction of the liquid biopsies (n=180, 19.1%), including all 10 control subjects with no clinical findings (i.e., no pathologies detected on EGD) along with peptic diseases (n=51), premalignant lesions (n=55), and gastric cancer cases (n=64), we evaluated the proportion of human- versus bacteria-derived DNA in gastric fluids by using quantitative PCR (Albuquerque et al., 2022).

### Pathological analysis of inflammatory cell infiltrates in tissue biopsies

Tissue biopsies were available from a subset of liquid biopsies evaluated for the gfDNA content, allowing the investigation of inflammatory cell infiltrates. In such cases, pathological tissue analyses were done independently by two expert cancer pathologists, both specialized in upper gastrointestinal tract neoplasia, who were blinded to the gfDNA concentration results from the liquid biopsies. Discrepant cases for the immune infiltration levels (n=3) were revised and discussed, and a consensus was achieved. After the microscopic identification of tumor cells and the histological classification of gastric cancer, the presence and intensity of inflammatory cell infiltrates in the neoplastic lesions were also evaluated, as well as their patterns in the tumor stroma (e.g., diffuse, microscopic lymphoid aggregates, lymphoid aggregates with the formation of hyperplastic germinal centers). We have set the predominant cellular composition of the inflammatory infiltrates, and quantified the infiltrate/tumor stroma ratios as follows: (i) minimal/rare – rare inflammatory cells in the tumor stroma; (ii) moderate – intermediate cellularity, no distortion of the epithelial component and without formation of luminal micro-abscesses, and (iii) intense/elevated – large amounts of inflammatory cells, making it difficult to visualize the stroma, with the formation of luminal micro-abscesses and/or epithelial thinning. We have also evaluated the presence of lymphoid aggregates, necrosis/ulcer and neutrophilic infiltrates. Areas directly related to ulcer formation and/or granulation tissue were not considered in the polymorphonuclear component assessment.

### Statistical analysis

Baseline gfDNA values for patient groups are given as a mean. Analyses were performed in the R statistical environment (version 4.2.0), as described (R Core Team, 2022). Statistical analyses were carried out by using one-way ANOVA, Tukey’s HSD, Mann-Whitney test and the Kruskal-Wallis test for paired or multiple comparisons. Survival data were collected for all subjects and the cutoffs on gfDNA concentrations, determined to separate survival groups, were calculated using the log-rank test (function maxstat.test from R package maxstat; Hothorn, 2017). Distributions of gfDNA concentrations for the four groups of patients (i.e., normal, peptic ulcer diseases, preneoplastic diseases, and gastric cancer) were compared by using the two samples Wilcoxon test, and p-values were adjusted by the Benjamini-Hochberg procedure (i.e., False Discovery Ratio, FDR). Logistic Regression Analysis was used to determine Receiver Operating Characteristics (ROC) and Area Under the Curve (AUC). Results were considered statistically significant when p ≤ 0.05.

## Results

### Patient groups and diagnoses

Gastric fluids were collected and gfDNA was extracted from a total of 1,056 subjects. After excluding 115 cases with diagnosis of hepatic portal hypertension, partial or total gastrectomy, esophagectomy, or other non-gastric cancer malignancies (e.g., esophageal squamous cell carcinoma, duodenal adenocarcinoma, Hodgkin’s disease, well-differentiated neuroendocrine tumor, and metastatic breast cancer, among others), a total of 941 patients (89.1%) remained for gfDNA analysis. These patients were grouped according to the EGD findings as: normal EGD (n = 10, 1.1%), non-malignant peptic diseases (n = 596, 63.3%) including erosive gastritis, erosive and non-erosive esophagitis with or without hiatal hernia, infectious or inflammatory disorders), preneoplastic conditions (n = 99, 10.5%) including atrophic gastritis, gastric intestinal metaplasia, dysplasia, Barret’s esophagus, or gastric adenocarcinoma (n = 236, 25.1%).

Diagnostic group, age, sex, and body-mass index (BMI) of the participants are shown in **Table 1**.

**Table 1.** Clinical and demographic attributes of participating subjects (n=941)

|  |  | **Non-gastric cancer\***<br>**(n=705, 74.9%)** | **Gastric cancer**<br>**(n=236, 25.1%)** |
| --- | --- | --- | --- |
| **Age** | ≤45 | 130 (18.4%) | 34 (14.4%) |
|  | 45-60 | 224 (31.8%) | 83 (35.2%) |
|  | ≥60 | 343 (48.7%) | 119 (50.4%) |
|  | Missing data | 8 (1.1%) | 0 |
| **Sex** | Male | 330 (46.8%) | 150 (63.6%) |
|  | Female | 367 (52.1%) | 86 (36.4%) |
|  | Missing data | 8 (1.1%) | 0 |
| **\*\*BMI** | Underweight | 10 (1.4%) | 9 (3.8%) |
|  | Normal weight | 206 (29.2%) | 91 (38.6%) |
|  | Overweight | 310 (44%) | 84 (35.6%) |
|  | Obese | 177 (25.1%) | 51 (21.6%) |
|  | Missing data | 2 (0.3%) | 1 (0.4%) |

\* Includes all patients diagnosed as presenting normal mucosa, with peptic diseases or pre-neoplastic lesions.
\*\*Body-mass index (BMI): Underweight, BMI<18.5; Normal weight, BMI=18.5-24.9; Overweight, BMI=25-29.9; Obese, BMI≥30

### Gastric fluid DNA concentrations according to age, pH and body-mass index

No significant variations in gfDNA concentration were found by age, pH of gastric fluid, or BMI when examined by group amongst gastric cancer patients or non-cancer controls. Regarding age, subjects were clustered in the three groups (<45, 45-60, and >60 years old) and no significant differences were found between subjects with (p = 0.26) or without (p = 0.14) gastric cancer. Similarly, no difference was found by pH analysis of gastric fluid (grouped as pH<7, pH=7, or pH>7); we found p-values of 0.60 and 0.90 respectively for gastric cancer and non-cancer controls. For BMI analysis, subjects were grouped as underweight (BMI< 18.5), normal (BMI between 18.5 - 24.9), overweight (BMI between 25 - 29.9), or obese (BMI ≥ 30). Again, no differences in gfDNA concentrations were found by BMI in individuals with gastric cancer (p = 0.59) or non-cancer controls (p = 0.25).

### gfDNA concentrations and sex

Next, we evaluated whether gfDNA concentrations vary according to sex in patients with gastric cancer and subjects without cancer. For the non-cancer cohort, we observed elevated gfDNA concentrations in females versus males (p = 7.44e^-4^), but no differences were seen between males and females in the cancer group (p = 0.68) (**Figure 1**). Therefore, when we compared gfDNA concentration between cancer versus non-cancer subjects, considering sex, we found striking differences for men and for women, respectively, p = 8.75e^-12^ and p = 1.20e^-12^ (**Figure 1**).

**Figure 1.**
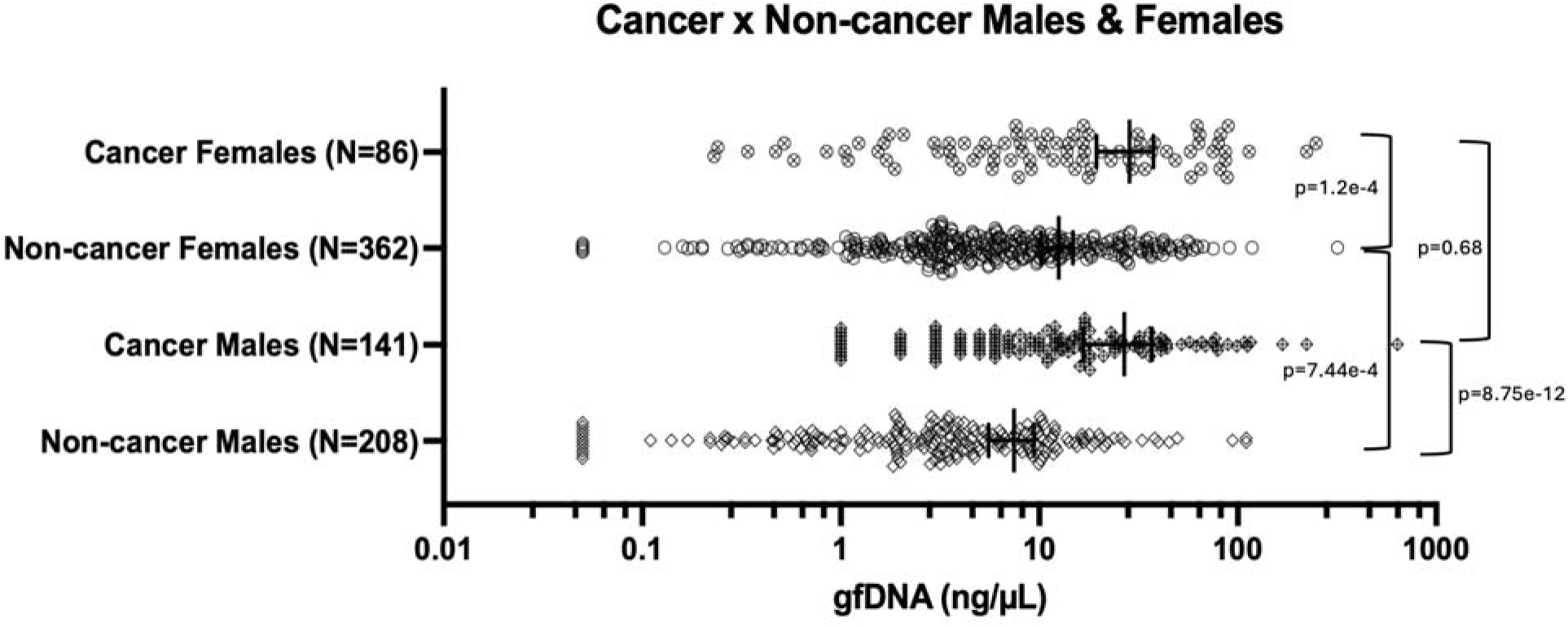
gfDNA concentrations observed in all 941 subjects, grouped according to sex, and presence of gastric cancer. Non-cancer subjects have normal gastric mucosa or minor peptic diseases (excludes pre-neoplastic disease). Bars indicate mean gfDNA concentration (ng/µL) and 95% confidence intervals.

As some studies have suggested that the use of proton pump inhibitors (PPI) is more frequent in women (Linseisen et al., 2022), we evaluated possible links between gfDNA concentrations and the reported history of current, previous, or no PPI use. In general, we saw no impact of PP on gfDNA concentrations. However, for the group of non-cancer subjects that used PPI (n=410), increased gfDNA concentrations were seen for women (n=206), as compared to men (n=204) (p= 1.51e^-4^), apparently reflecting the sex-related results shown in **Figure 1**, between males and females, unrelated to PPI-use. Moreover, gfDNA concentrations were not different for subjects that reported current and previous administration, as well as no PPI administration, for females with or without cancer (n=55, n=6 and n=25; n=111, n=45 and n=206, respectively), as well as for males with or without cancer diagnosis (n=60, n=19 and n=62; n=75, n=51 and n=209, respectively, with corresponding p-values of p= 0.92; p= 0.07; p= 0.24 and p= 0.71).

### Analysis of bacteria to human ratios in gfDNA

As the gfDNA origin is likely to be a sum of human DNA (including epithelial, stromal, and immune cells, tumor and non-tumor cells), combined with microbiota-derived DNA (transient, saliva-derived microorganisms, the resident microbiota of the stomach and other upper digestive tract structures), we performed a quantitative analysis of bacteria and human DNA for a subset of samples (n=180), based on human-to-bacteria DNA ratios, as recently described (Albuquerque et al., 2022). The samples selected for the analysis of this subset analysis included all 10 controls without any pathologies detected, as well as samples from the peptic diseases group (n=51), preneoplastic conditions (n=55) and the gastric cancer group (n=64). This analysis revealed no significant differences in the ratios of human- or bacteria-derived DNA found in the gfDNA (**Suppl. Table 1**).

### gfDNA concentrations are increased in gastric cancer patients compared to non-cancer controls

We first determined the concentration of gfDNA from subjects with different diagnoses after EGD and found the following values for the groups: normal mucosa and peptic diseases – mean 10.77 ng/µL; 95% CI: 9.23 to 12.33; n=606; preneoplastic conditions - mean 10.10 ng/µL; 95% CI: 7.59 to 12.60; n=99 and gastric cancer - mean 26.86 ng/µL; 95% CI: 20.05 to 33.79; n=236. Notably, mean concentration of gfDNA was significantly different among the groups (p=3.61e^-12^). gfDNA concentration was higher in gastric cancer as compared to the combined group of normal EGD and peptic disease patients (p=9.55e^-13^), as well as to the preneoplastic group (p=1.10e^-5^). In contrast, gfDNA concentrations were not different between the non-cancer groups (p=0.89) (**Figure 2A**).

**Figure 2.**
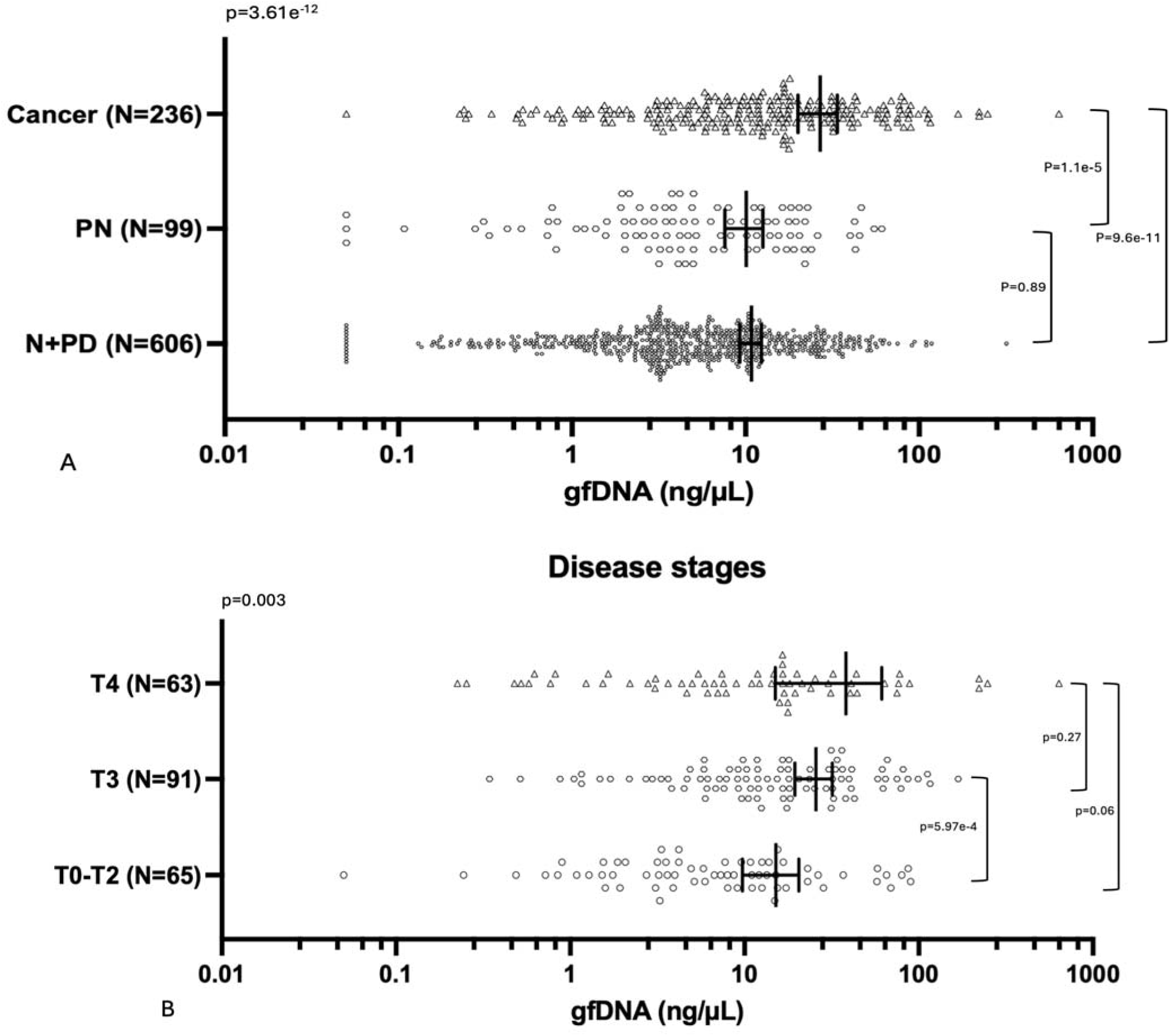
gfDNA concentration according to disease diagnosis and tumor stage. A) gfDNA concentration (ng/µL) in patients with no endoscopic findings: Normal (N) or presenting minor peptic diseases (PD) – N+PD; preneoplastic conditions (PN), or gastric cancer (Cancer). B) gfDNA concentrations (ng/µL) for early-stage disease patients – T0-T2 (T0 + Tis + T1 + T2), as compared to more advanced disease stages T3 and T4. Bars indicate mean gfDNA concentration (ng/µL) and 95% confidence intervals.

For gastric cancer patients, we evaluated possible correlations between gfDNA concentrations and clinicopathological variables such as tumor-stage and histopathological grades. In general, increments in gfDNA were found in subjects as the disease progresses (**Figure 2B**). Notably, the gfDNA mean concentration for early-stage disease (tumor stages - T0 + Tis + T1 + T2, or T0-T2 n=65) was 15.12 ng/µL; 95% CI: 9.73 to 20.50, increasing in T3 (25.66 ng/µL; 95% CI: 19.46 to 31.85; n=91; p = 5.97e^-4^) and T4 stages (38.12 ng/µL; 95% CI: 15.02 to 61.22; n=63; p =0.06).

Other comparisons were not significant, including gfDNA concentrations for patients with localized (n=165) versus metastatic disease (n=71). gfDNA concentrations could not differentiate tumor histopathological grades (p>0.05), or Lauren gastric cancer subtypes: diffuse (n=97), intestinal (n=93), or mixed (n=25) (p= 0.28) (data not shown).

### gfDNA correlates with the presence of inflammatory cells infiltrates and gastric cancer diagnosis and survival

Our liquid biopsy results revealed increased gfDNA concentrations in gastric cancer patients as compared to non-cancer controls. To explore whether gfDNA concentrations could be used to support gastric cancer diagnosis, we analyzed the ROC curve of gfDNA between gastric cancer versus non-cancer controls. We observed a statistically significant curve (AUC=0.66; p=5.23e^-10^), with a fair capability of detecting true positive gastric cancer patients by simply considering gfDNA concentrations (**Suppl. Figure 1A; Suppl. Table 1**), indicating its fair diagnostic support value. We also found a statistically significant curve of gfDNA between cancer/preneoplastic conditions and peptic diseases/normal (AUC=0.61; p = 6.86e^-7^) (**Suppl. Figure 1B**).

We next evaluated if gfDNA concentrations would correlate with disease prognosis. Well-annotated clinical records were collected for all subjects to determine gfDNA concentrations cutoff values that would split the patients into groups with contrasting overall survival through a log-rank test. By using this approach, a gfDNA cutoff of 1.28 ng/µL was set and gastric cancer patients presenting gfDNA concentrations above this threshold had better overall survival (n=163; p = 0.019), even more significant after patients with metastatic tumors were not considered (n=148; p = 0.014) (**Figure 3A**). Finally, to further investigate the possible link between patient survival and gfDNA concentration, we examined a subset of gastric cancer patients (n=32), for which pathology slides were available, to investigate possible correlations between gfDNA concentrations and the intensity of inflammatory cell infiltrates (**Figure 3B**). For this we managed to include 8 samples with gfDNA below the threshold, and 24 above it. Even with a small sample set, this result was also significant (p=0.001) (**Figure 3C and Suppl. Table 3**), suggesting a possible mechanism to support the correlation of gfDNA concentration and gastric cancer survival data (**Figure 3A**).

**Figure 3.**
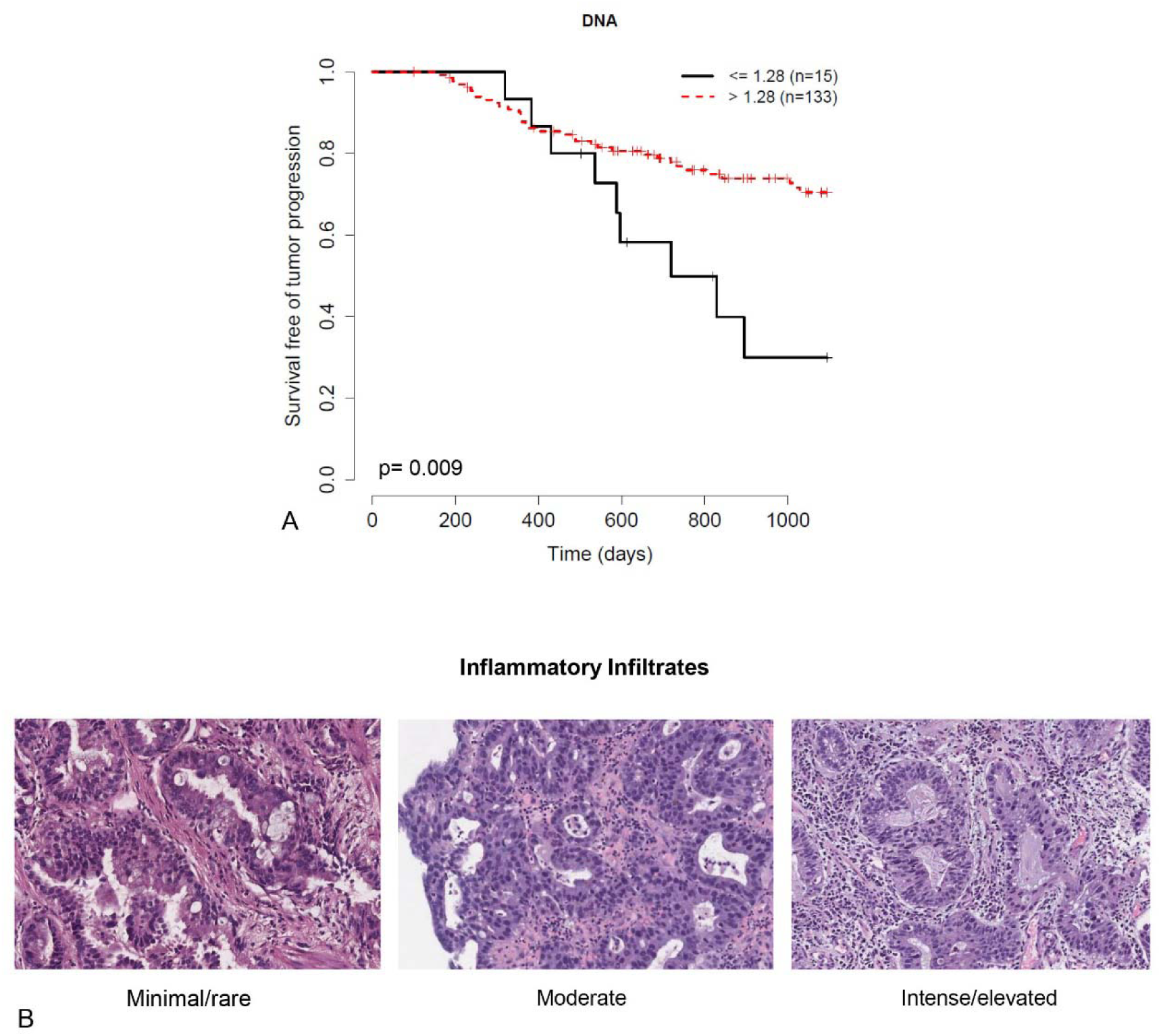
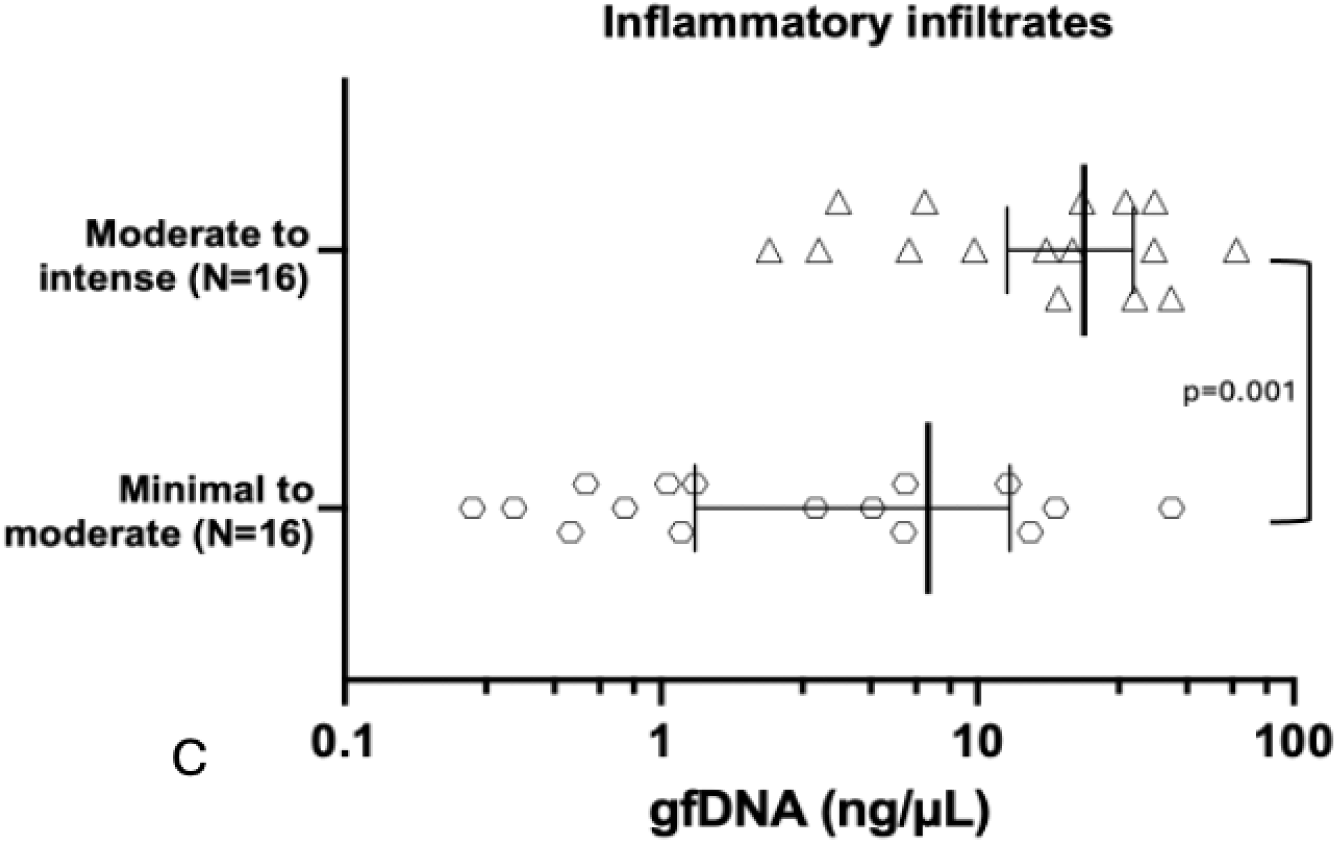
gfDNA analysis according to patient survival and infiltration of inflammatory cells. (A) For a total of 148 non-metastatic GC patients, cutoff gfDNA concentrations determined by a log-rank test (1.28 ng/µL) discriminate survival free of tumor progression (p = 0.009). (B) For a subset of 32 cases, for which representative biopsy H& slides were available, inflammatory cell infiltrates in the gastric tumors were inspected by experienced pathologists and (C) have shown that more intense inflammatory infiltrates in tumors are characteristic of subjects with higher gfDNA concentrations (p = 0.001). Bars indicate mean gfDNA concentration (ng/µL) and 95% confidence intervals.

## Discussion

Gastric cancer remains a leading cause of tumor mortality worldwide, a clinical feature attributable at least in part to its often-late stage at diagnosis and/or lack of prognostic factors to enable personalized disease management. However, with varying access to subspecialists a role for gfDNA may help in the clinical management of patients. Early gastric cancer detection may often be delayed during EGD if biopsies samples are either taken too superficially or if the tissue is too scanty for meaningful pathological analysis. Thus, the translational application of a biomarker that provides support to gastric cancer diagnosis and prognosis would be useful, especially in a less specialized outpatient ambulatory or inpatient hospital settings.

In 1977, Leon et al. published a seminal paper showing that cancer patients had higher levels of cell-free DNA (cfDNA) in their blood compared to healthy individuals, suggesting its use as a potential cancer biomarker. Since then, numerous studies confirmed these findings and have explored the potential of using cfDNA for cancer diagnosis, prognosis, and monitoring treatment response, including gastric cancer. Qian et al.(2017) analyzed cfDNA in serum samples of subjects with gastric dysfunctions, including gastric cancer, and found that cancer patients presented elevated cfDNA when compared to benign gastric disease or controls.

However, the diluted amounts of tumor-derived/tumor-microenvironment DNA remains minute in the peripheral circulation. This prompted us to determine DNA concentrations in the gastric fluids of subjects undergoing EGD to investigate non-specific digestive symptoms.

Patients included those with diverse peptic diseases (controls), pre-neoplastic lesions as well patients with confirmed gastric cancer. We reasoned that liquid biopsies of gastric fluid incidentally obtained during EGD would offer diagnostic support and would possibly offer clinically useful information. Whereas EGD is an invasive procedure, most of these patients will receive at least one diagnostic EGD during their initial workup and tumor staging, and gastric fluids collected at this moment can be useful. However, prior studies of gastric fluids have heretofore been limited to the evaluation of certain molecules, such as proteins, RNAs or for the analysis of mutations in specific genes (Wu, Chung, 2013; Shao et al., 2014; Pizzi et al., 2019). Here we show that the simple evaluation of gfDNA concentration serves as a convenient and promising biomarker to support gastric cancer diagnosis and staging and correlates with immune cell infiltrates. In a large cohort, we show that increased DNA concentrations in gastric fluids may differentiate gastric cancer patients from individuals without cancer, independent of age, BMI, sex, PPI use, or Lauren’s histological subtypes. Increasing amounts of gfDNA were significantly associated with gastric cancer diagnosis, as well as advanced tumor stages.

Of note, there were several interesting observations from our results. Overall, we found that T4 tumors had gfDNA concentration similar to T3 tumors (T3: 25.66 ng/µL; 95% CI: 19.46 to 31.85; and T4: 38.12 ng/µL; 95% CI: 15.02 to 61.22; p=0.27) and the difference between T4 and earlier stages (T2 and below) was non-significant (p = 0.065). One might speculate that this may be due to the higher variability of gfDNA concentration seen for T4 (**Figure 2B**) that could be intrinsic to the T4 stage, as some tumors are more invasive than others, and if the tumor grows through the gastric wall and the serosa (Wu, Chung, 2013) it might perhaps be shedding DNA into adjacent peritoneal structures instead of just the gastric cavity, leading to a reduction of tumor-derived DNA in the gastric cavity. This contrasts with T0-T3 tumors that appear to continuously and preferentially release DNA inside the gastric cavity. In agreement with this possibility, we found that advanced tumors have increased gfDNA concentration as compared to early stages, a finding that might suggest that the origin of the gfDNA could perhaps derive from the tumor, but also from the tumor environment, from immune cell infiltrates, and the microbiota. However, our quantitative analysis of bacterial and human DNA in the gfDNA for a set of samples (n=180), including gastric cancer samples (n=64), showed no clear trends towards an increased bacterial DNA content, rendering that possibility unlikely (**Suppl. Table 2**). We also observed an intriguing correlation between gfDNA concentration and survival, with gastric cancer patients presenting lower gfDNA concentrations (set at ≤ 1.28 ng/µL) associated with a worse prognosis. We envisaged that this observation could be associated with poorer immune responses (immunologically “cold tumors”), which was supported by the finding that patients with higher gfDNA concentrations (set at >1.28 ng/µL) more often presented with moderate-to-intense inflammatory infiltrates, whereas individuals with lower gfDNA usually presented minimal-to-moderate inflammatory infiltrates (p=0.001). Notably, two-thirds (16 out of 24) of the cases with increased gfDNA had moderate/intense/elevated immune cells infiltrates and all cases within this classification of inflammatory infiltrates were in the group of increased gfDNA (**Suppl. Table 3**). This suggests that the elevated gfDNA concentration observed in subjects with better outcomes may originate from two primary sources: an active population of infiltrating immune cells, together with the tumor cells eliminated by these immune infiltrates.

The results presented here suggest wide implications for the translational application of liquid biopsies with gfDNA concentration analysis in the diagnosis, staging and management gastric cancer patients. The most obvious barrier to the translational application of liquid biopsies and gfDNA measurement in the routine clinical setting is the need of an EGD to access it. However, the use of gfDNA may hold the highest benefit by increasing the value of information obtained from the initial EGD, which is --as mentioned previously--a procedure that all patients with gastric cancer will eventually undergo multiple times during their disease workup. Indeed, the suction of gastric fluids during endoscopy is readily performed during EGD, when the gastric walls must be free of fluids for a better visualization of the mucosa. Thus, the convenient incorporation of this liquid biopsy approach during EGD is cost-effective and a minor additional step to the procedure. The benefit is that the continuous proximity of gastric fluids to the tumor site portends a much-reduced dilution of gfDNA, some of it being tumor-derived DNA, which allows a better representation of tumor-related DNA alterations. In a comparative sense, liquid biopsy of gastric fluid with gfDNA measurement is similar to obtaining DNA from peritoneal lavage of patients with endometrial cancer, but indeed much harder to obtain, in which peritoneal fluid has higher mutant allelic fractions as compared to plasma (Mayo-de-las-Casas, 2020), similar to previous finding in gastric cancer in our early work (Pizzi et al., 2019).

Much interest has been given to developing new tools for detecting diagnostic, prognostic, and predictive markers in gastric cancer (Mayo-de-las-Casas, 2020). The area under the receiver operating characteristic (AUROC) analysis for liquid biopsy of gastric fluid with gfDNA analysis yielded a discrimination capability of 0.66, which is comparable to serological tumor biomarkers such as CEA (0.68), CA72-4 (0.67), or CA19-9 (0.64) that are currently used in standard oncological practice (Tahara, Arisawa, 2015; Yu et al., 2016; Lin et al., 2020). Moreover, the monitoring of gfDNA concentration may also be valuable for a closer follow up and better diagnostic accuracy of patients with potentially premalignant diseases and/or at higher genetic/familial risk of developing gastric cancer. One might also speculate that our findings could perhaps also apply to other upper gastrointestinal malignant tumors such as esophageal cancer and/or gastroesophageal junction neoplasia, and future studies should rigorously evaluate those open possibilities.

This study represents an initial evaluation of the prognostic impact of gfDNA concentration in human subjects diagnosed with gastric cancer, as compared to other conditions such as pre-malignant lesions and peptic diseases. The participants were referred to diagnostic EGD and their distribution in the diagnostic groups reflect the real-life case numbers seen in a single institution, a dedicated cancer center. Before receiving sedation for the EGD, participants were invited to join the study. Therefore, samples were collected prospectively, before diagnosis, from all subjects and no attempts to balance the groups were made to avoid introducing any potential selection bias. Limitations also include the primer sets that allow a partial quantification of some bacterial groups (i.e., microbiome analysis), the limited number of pathology slides available from gastric cancer patients to investigate immune cell infiltrates (n=32), and the lack of a more comprehensive approach that would also allow a cell type-specific deconvolution and a better explanation of the functional mechanisms that lead to increased gfDNA concentrations. The diagnostic potential of elevated gfDNA levels was not explored together with other known biomarkers such as CEA or CA19-9. Our data suggest that gfDNA is not only derived from tumor cells but may have expressive amounts of DNA derived from infiltrating immune cells. The precise determination of the presumed cell-of-origin of this increased gfDNA remains to be unequivocally demonstrated as it would have to rely on methods not used in this study, such as RNA-Seq and DNA-seq.

Moreover, the large patient cohort analyzed in this study was derived from a single specialized institution (a tertiary cancer center), and it is therefore unclear whether our findings might be broadly generalized; this open question should be addressed in further confirmatory studies from other hospital types and geographical regions.

Finally, for considering subjects with an established gastric cancer diagnosis, gfDNA has the potential to serve as a rapid, low-cost surrogate marker of the tumor immune microenvironment, whose characterization may have implications for predicting chemotherapy and immunotherapy response (Jiang et al., 2018; Jiang et al., 2019; Chen et al., 2022; Espinosa-Carrasco et al., 2024), and prognostic repercussions that, in the future, could be useful to treatment planning and risk-adapt therapeutic strategies. As we have gathered no data on gfDNA changes over time, conclusions on its utility for monitoring treatment response and/or predicting recurrence are currently limited, but warrant further investigation.

Given that gastric cancer has currently no suitable tumor prognostic biomarker, one hopes that the initial results of this report will encourage the liquid biopsy of EGD-collected gastric fluids to evaluate gfDNA concentrations towards clinic-ready translational applications. Further gastric fluid studies may help revolutionize gastric cancer care especially in places where specialties may not be readily available. Also, it may assist as a first line exam that can be done to help detect cancer cases, with prognostic value. Whereas more research is needed, early data indicates this may be a way to provide access and availability for primary and recurrent gastric cancer.

## Author Contributions

Collected samples and clinical data: FCC, AGP, CZS, LBCS, GPB, FAP, LLSA

Concepted and performed analysis: FCC, TFB, AD, RD, ITS, DNN, EDN

Pathology: APB, GOS, WAN

Data analysis and statistics: FCC, TFB, AD, RDD, ITS, RP, WA, DNN, EDN

Discussion of findings: FCC, TFB, AGP, CZS, LGVC, ML, HI, SL, HSW, FJFV, ITS, RC, RP, WA, DNN, EDN

Writing & editing: FCC, TFB, AGP, CZS, LGVC, ML, HI, SL, HSW, FJFV, ITS, RC, RP, WA, DNN, EDN

Final manuscript review and approval: All authors

Project supervision: EDN

## Data Availability

All data produced in the present study are contained in the manuscript and in its supplementary information files

## Acknowledgements

Authors acknowledge the support given by the institutional Tumor Bank from A. C. Camargo Cancer Center. We thank Dr. Christian Abnet and Dr. M. Constanza Camargo (NCI, NIH, USA) for their critical review of this manuscript.

## Cadoná et al., Supplementary Information

**Supplementary Figure 1.**
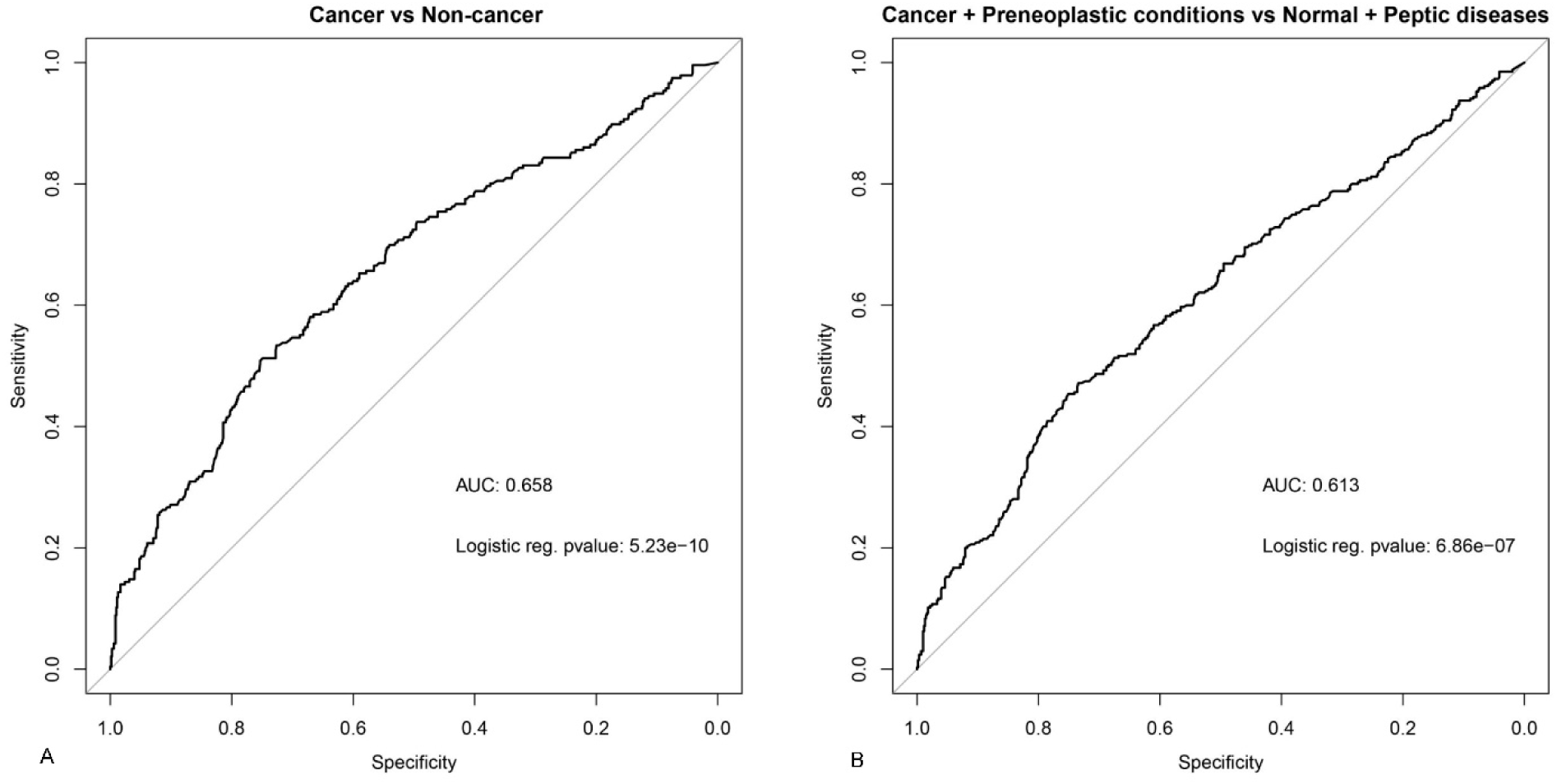
Analysis of Receiver Operating Characteristics (ROC) and Area Under the Curve (AUC) for gfDNA levels between cancer patients and non-cancer individuals. A) ROC curve of gfDNA between the cancer patients and non-cancer individuals. B) ROC curve of gfDNA between cancer versus non-cancer and cancer + preneoplastic conditions versus peptic diseases + normal.

**Supplementary Table 1.** Performance metrics of the ROC/AUC curves shown in Suppl. Figure 1, panels A and B.

| **Cancer vs. non-cancer thresholds**<br>**(Suppl. Fig. 1A)** |  |  |  | **Cancer + pre-neoplastic lesions vs. normal +**<br>**peptic diseases (Suppl. Fig. 1B)** |  |  |  |
| --- | --- | --- | --- | --- | --- | --- | --- |
| **Threshold** | **Specificity** | **Sensitivity** | **Youden_J** | **Threshold** | **Specificity** | **Sensitivity** | **Youden_J** |
| 11.81 | 0.7532 | 0.5085 | 0.2617 | 10.75 | 0.7343 | 0.4716 | 0.2060 |
| 11.75 | 0.7489 | 0.5127 | 0.2616 | 11.55 | 0.7508 | 0.4537 | 0.2046 |
| 11.65 | 0.7475 | 0.5127 | 0.2602 | 10.65 | 0.7310 | 0.4716 | 0.2027 |
| 10.75 | 0.7262 | 0.5339 | 0.2601 | 11.65 | 0.7541 | 0.4478 | 0.2019 |
| 11.86 | 0.7532 | 0.5042 | 0.2574 | 10.9 | 0.7360 | 0.4657 | 0.2016 |
| 10.65 | 0.7234 | 0.5339 | 0.2573 | 11.485 | 0.7475 | 0.4537 | 0.2013 |
| 10.55 | 0.7220 | 0.5339 | 0.2559 | 10.55 | 0.7294 | 0.4716 | 0.2010 |
| 11.55 | 0.7418 | 0.5127 | 0.2546 | 11.81 | 0.7591 | 0.4418 | 0.2009 |
| 10.455 | 0.7163 | 0.5381 | 0.2544 | 11.465 | 0.7459 | 0.4537 | 0.1996 |
| 10.9 | 0.7277 | 0.5254 | 0.2531 | 11.75 | 0.7541 | 0.4448 | 0.1989 |

**Supplementary Table 2.** Analysis of CT values for bacteria and human target genes.

|  | n | Median CT | p |
| --- | --- | --- | --- |
| CT 16S-V1 |  |  |  |
| Normal | 10 | 17.27 | 0.21 |
| Peptic diseases | 51 | 18.60 |  |
| Preneoplastic conditions | 55 | 17.72 |  |
| Cancer | 64 | 18.92 |  |
| CT β-ACT |  |  |  |
| Normal | 10 | 25.29 | 0.07 |
| Peptic diseases | 51 | 26.21 |  |
| Preneoplastic conditions | 55 | 27.01 |  |
| Cancer | 64 | 27.46 |  |
| Ratio 16S-V1/β-ACT |  |  |  |
| Normal | 10 | 0.68 | 0.83 |
| Peptic diseases | 51 | 0.71 |  |
| Preneoplastic conditions | 55 | 0.66 |  |
| Cancer | 64 | 0.69 |  |

**Supplementary Table 3.** gfDNA concentration and intensity of inflammatory cell infiltrates.

| **Inflammatory infiltrates** | **Lower gfDNA (<1.28ng/μL)**<br>**N=8 patients** | **Higher gfDNA (>1.28ng/μL)**<br>**N=24 patients** |
| --- | --- | --- |
| **Minimal/rare** | 0 | 8.3% (n=2) |
| **Minimal/rare to moderate** | 62.5% (n=5) | 12.5% (n=3) |
| **Moderate** | 37.5% (n=3) | 12.5% (n=3) |
| **Moderate to intense/elevated** | 0 | 50% (n=12) |
| **Intense/elevated** | 0 | 16.6% (n=4) |

